# Effects of short-term restriction of animal products on blood biomarkers and cardiovascular disease risk

**DOI:** 10.1101/2023.05.17.23290094

**Authors:** Eleni M. Loizidou, Alexandros Simistiras, Petros Barmpounakis, Stavros Glentis, Alexandros Dimopoulos, Maria Anezaki, Ioannis Kontoyiannis, Nikolaos Demiris, Pavlos Rouskas, Nikolaos Scarmeas, Mary Yannakoulia, Konstantinos Rouskas, Antigone S. Dimas

**Affiliations:** Institute for Bioinnovation, Biomedical Sciences Research Center ‘Alexander Fleming’, Fleming 34, 16672, Vari, Greece; biobank.cy Center of Excellence in Biobanking and Biomedical Research, University of Cyprus, Cyprus; Department of Statistics, Athens University of Economics and Business, Athens, Greece; Pediatric Hematology/Oncology Unit (POHemU), First Department of Pediatrics, University of Athens, Aghia Sophia Children’s Hospital, Levadias 8, 11527, Athens, Greece; Hellenic Naval Academy, Hatzikyriakou Avenue, Pireaus 185 39, Greece; Statistical Laboratory, Centre for Mathematical Sciences, University of Cambridge, United Kingdom; 1st Department of Cardiology, AHEPA University Hospital, Aristotle University, Thessaloniki, Greece; Department of Neurology, Columbia University, New York, NY, USA; 1^st^ Department of Neurology, Medical School, National and Kapodistrian University of Athens; Department of Nutrition and Dietetics, Harokopio University, Athens, Greece; Institute of Applied Biosciences, Centre for Research & Technology Hellas, Thessaloniki, Greece

**Author notes:** Corresponding author: Email address, Institute for Bioinnovation, Biomedical Sciences Research Center ‘Alexander Fleming’, Fleming 34, 16672, Vari, Greece.

**Keywords:** dietary restriction, biomarkers, disease prevention, inflammation

## Abstract

Dietary interventions constitute a means of untapped potential for the prevention and management of diseases including cardiovascular disease (CVD) and inflammatory disorders. The extent to which dietary modification can contribute to disease prevention and treatment however remains unclear. Here, we addressed the effects of a dietary pattern, involving periodic animal product restriction (APR), on markers of human health. We compared blood biomarkers, complete blood counts, anthropometric traits and blood pressure between two dietary states for a unique group of 200 individuals from Greece, who alternate between omnivory and APR, for religious reasons. We used linear mixed effects models to report changes in measured traits between dietary states. Short-term APR was linked to a reduction in levels of total and LDL cholesterol [both β=-0.3, p<0.0001], to a 72% decrease in CRP concentration [β=-1.3, p=0.001], to reduced platelet counts [β=-9.8, p<0.0001] and to a concurrent reduction in CVD risk (5.3% to 4.9%, p=0.02), shifting APR-practicing individuals from a high to a low-to-moderate risk group. Beneficial changes were recorded for markers of renal and liver function, with the exception of ALP [β=2.5, p<0.0001]. These changes were not detected in a control group of 211 continuously omnivorous individuals. Overall, short-term APR was linked to a biomarker profile associated with positive effects on health, suggesting that even modest dietary modifications can be harnessed towards disease prevention.

## Introduction

Dietary patterns that partially or wholly exclude animal products have been associated with a low risk of major chronic diseases, including cardiovascular disease, type 2 diabetes and inflammatory bowel disease ^1-3^. Indeed, there is growing evidence that plant-based diets have the potential to act as powerful interventions for the prevention and treatment of a wide range of conditions ^4^ with cross-sectional and interventional studies linking them to beneficial effects on BMI, total and LDL cholesterol, blood glucose, CRP, serum urea, urinary creatinine and γ-GT ^3,5-10^. Vegan diets in particular, which involve abstinence from all animal products, have been additionally associated with lower risk of cancer incidence, diabetes, diverticular disease, cataracts ^3,9,10^, but to a higher risk of bone fractures ^9-11^. Emerging evidence also suggests an impact on the immune system ^12,13^ with vegan diets being linked to improvement of symptoms in rheumatoid arthritis patients ^14^. Despite their positive impact on health, these diets can be too restrictive for practicing individuals, thus, individuals often choose to abstain only from some animal products or to reduce meat consumption. Consequently, there is a growing trend toward flexitarian approaches (flexible diets involving the consumption of a range of plant-based foods with limited consumption of animal products) which have been linked to health benefits including diabetes risk and blood pressure reduction ^15^. This dietary shift is also recommended by the European Society of Cardiology (ESC) with its latest guidelines recommending the substitution of meat with plant foods for the improvement of cardiovascular health ^16^.

Given their substantial beneficial effects, it is important to gain a comprehensive understanding of how dietary patterns that restrict animal products affect markers of human health. To date, most studies involving large numbers of individuals are cross-sectional and while they build on existing knowledge, they do not allow for causal inference and usually report on a relatively limited number of outcomes or measurements. Important insight has been provided by meta-analyses of observational studies and randomised controlled trials (RCTs), but these studies suffer from limitations arising from the heterogeneity of primary studies analysed. Heterogeneity pertains to the variability in dietary patterns (e.g. vegan, vegetarian, flexitarian), to the health status of participants (e.g. healthy, obese, at-risk, patients), to the dietary intake of control groups (e.g. no dietary intervention, specific dietary interventions), to lack of adjustment for important confounders (e.g. sex, age, and BMI) and to the duration of the dietary intervention (a few weeks to several months). Typically, most studies involve interventions of over 12 weeks and thus do not map acute responses to dietary interventions, although some studies involve shorter interventions ^12^.

To address the gaps in our knowledge, we established FastBio (Fasting Biology), a follow-up study focusing on a unique subgroup of the Greek population that follows a lifelong dietary pattern involving periodic restriction of animal products ^17^. These individuals alternate between omnivory and animal product restriction (APR), for religious reasons, in a highly structured manner. We explored effects of this dietary pattern on markers of human health and compared findings to those from a continuously omnivorous group from the general population.

## Methods

### Recruitment and selection of participants

The FastBio study was advertised during Autumn 2017 to the local communities of Thessaloniki, Greece through various means, including the press, radio, social media, and word of mouth. Candidate participants expressed their interest via telephone, email or via the project’s website (www.fastbio.gr). Following screening of over 1,000 candidates through two interviews, 411 apparently healthy, unrelated individuals, who met FastBio selection criteria (Supplementary Text S1) were included in the study. Participants belonged to one of two groups, specified by their dietary habits: 200 individuals who follow a temporally structured dietary pattern of periodic restriction of animal products (periodically restricted, PR group) and 211 individuals who follow the diet of the general population (non-restricted group, NR group). PR individuals constitute a unique study group given their consistent and predictable adherence to a specific dietary pattern (Supplementary Text S2). Restriction is practiced for religious reasons, as specified by the Greek Orthodox Church and involves alternating between omnivory and restriction of animal products, including meat, fish, dairy products and eggs (but not molluscs and shellfish), for 180-200 days annually. This dietary pattern involves four extended periods of APR throughout the year, and on on Wednesdays and Fridays of each week (Supplementary Fig. S1). During periods of restriction, PR individuals also typically consume less alcohol. When not restricted, PR individuals follow an omnivorous type of diet. Only PR individuals who had practiced periodic APR for at least ten years were included in the FastBio study. Notably, very few participants were receiving dietary supplements (Supplementary Text S3). NR participants were continuously omnivorous individuals from the general population, who were not under any kind of special diet.

### Collection of data, of biological material and measurement of traits

FastBio participants were invited to two scheduled appointments at the Interbalkan Hospital of Thessaloniki. The first appointment took place in October-November 2017 (Timepoint 1, T1) and covered a period where all PR individuals had been on an omnivorous diet (excluding Wednesdays and Fridays) for a period of eight or nine weeks. The second appointment (Timepoint 2, T2) took place in March 2018, and covered a period during which PR individuals had abstained from meat, fish, dairy products and eggs for three to four weeks during the abstinence period prior to Easter (Lent). At T2, a recall rate of 95% was achieved, with 192 (out of 200) PR participants and 198 (out of 211) NR participants attending both appointments. All appointments were scheduled between 7:30-9:30 am to minimize circadian effects and were completed during a two-week window to minimize effects of seasonality ^18^. The ethics committee of BSRC Alexander Fleming approved the study protocols, the study was performed in line with the principles of the Declaration of Helsinki, and all participants provided written informed consent.

For each participant, following overnight fasting, a blood sample was collected by a certified nurse and was taken for biomarker and complete blood count (CBC) testing at the InterBalkan Hospital facilities (cobas c 311 analyzer, Roche Diagnostics). We report results on 16 blood (serum) biomarker traits and 23 CBC traits (Supplementary Table S1). Biomarkers were grouped to allow for broad interpretation ^5^: blood lipids (total cholesterol, LDL cholesterol, HDL cholesterol, triglycerides), glucose metabolism (glucose, insulin, HbA1c), renal function (urea, uric acid, creatinine), liver function (aspartate aminotransferase (AST), alanine aminotransferase (ALT), gamma-glutamyltransferase (γ-GT)), bone and liver function (alkaline phosphatase (ALP)), thyroid function (thyroid-stimulating hormone (TSH)), and inflammation (C-reactive protein (CRP)). Using glucose and insulin measurements, we calculated homeostatic model assessment (HOMA) indices to assess beta-cell function (HOMA-B) and insulin resistance (HOMA-IR) (Supplementary Text S4). Height was measured to the nearest 0.5 cm using a stadiometer. Body weight was measured with the use of a calibrated digital scale with an accuracy of ±100g. BMI was calculated as body weight divided by the square of height (kg/m^2^). Systolic and diastolic blood pressure (SBP and DBP respectively) were measured using a traditional sphygmomanometer.

## Statistical analysis

We performed multivariable regression analysis using linear mixed effects (lme) models with random effects (“nlme” R version 4.1.1) to report within-group changes in measured traits between T1 and T2, with T2 (restriction timepoint) as the reference timepoint (Supplementary Text S5). Between-group differences within each timepoint are also reported, with PR as the reference group (Supplementary Text S5). Tukey’s correction was performed for all associations and *P*-values < 0.05 were considered statistically significant. Prior to statistical analysis, the sampling distribution for each trait was investigated and eight non-normally distributed traits (triglycerides, glucose, insulin, creatinine, AST, ALT, γ-GT and CRP) were log-transformed prior to regression analyses. For CRP, 49 individuals (25 PR and 24 NR) with values suggesting acute inflammation (CRP>5 mg/L) were excluded from further analysis. All statistical analyses were performed using R software, version 4.1.1.

### Blood biomarkers and CBC traits

We fitted lme models on measured traits with sex, age, and BMI as fixed confounders (Supplementary Text S6, Table S2) and included an interaction term between dietary group (PR or NR) and timepoint (T1 or T2) to conduct comparisons. We also corrected for medication use to account for effects on measured traits (Supplementary Text S7). Adjusting for smoking and alcohol consumption did not have an impact on our results (Supplementary Text S7). Individual heterogeneity was accounted for by including participant ID as a random intercept.

### Anthropometric traits and blood pressure

We fitted lme models on weight and BMI as described above, but without adjustment for BMI. We also fitted lme models for SBP and DBP with the above covariates, adjusting additionally for smoking status, which is a known risk factor for hypertension ^19^. Participants were defined as smokers and non-smokers (with past smokers and e-cigarette users also considered as non-smokers) based on NHS guidelines (https://www.nhs.uk/live-well/quit-smoking/using-e-cigarettes-to-stop-smoking/).

### Calculation of 10-year CVD risk score

The 10-year CVD risk score was calculated for participants ≥ 40 years old, as specified by the ESC guidelines, through the SCORE2 risk calculators (https://u-prevent.com/calculators/ascvdScore). ESC defines four risk groups based on country-specific CVD mortality, with Greece belonging to the moderate risk group, therefore the moderate risk calculator was used ^20^. The SCORE2 and SCORE2-OP algorithms were applied to individuals aged 40-69 years old and ≥ 70 years old respectively. The scores take into account age, sex, smoking status, SBP, total cholesterol and HDL cholesterol as predictors and are applicable to apparently healthy individuals with no previous CVD or type 2 diabetes mellitus. We calculated the score for 158 PR individuals (89 females and 69 males) and 122 NR individuals (65 females, 57 males) for both timepoints. A Wilcoxon signed-rank test was used to assess the change (%) in the CVD risk score between timepoints and a *P*-value < 0.05 was considered statistically significant.

## Results

### Study population characteristics

Table 1 and Supplementary Table S2 show the characteristics of FastBio participants at T1. Compared to NR individuals, PR were on average older and had higher BMI and blood pressure (Table 1). Both groups consisted of slightly more female participants, while most individuals were non-smokers (never smoked or stopped) with PR individuals being less likely to smoke. The vast majority were apparently healthy (according to self-reports) although a minority had underlying chronic conditions, including diabetes, arterial hypertension and hypothyroidism, for which treatment was being received (Supplementary Table S3). Mean values of blood biomarkers and CBC traits at both timepoints are summarised in Supplementary Table S1.

**Table 1.** Study population characteristics at timepoint 1 (T1) ^a^.

|  | PR (N=200) | NR (N=211) | Total (N=411) |
| --- | --- | --- | --- |
| Age (years) | 51.5 ± 13.5 | 45.0 ± 13.1 | 48.1±13.6 |
| Sex (female %) | 108 (54.0) | 116 (55.0) | 224 (54.5) |
| BMI (kg/m<sup>2</sup>) | 28.4 ± 4.6 | 26.2 ± 4.4 | 27.3 ± 4.6 |
| Smoking status (Yes %) | 13 (6.5) | 70 (33.2) | 83 (20.2) |
| Blood pressure (mmHg) |  |  |  |
| SBP | 127±19.0 | 121±19.7 | 124±19.4 |
| DBP | 80±10.8 | 78±11.8 | 79±11.3 |

<sup>a</sup> Continuous variables were expressed as mean ± standard deviations and categorical variables as %.
BMI = Body Mass Index, SBP = Systolic Blood Pressure, DBP = Diastolic Blood Pressure

### Within-group changes in measured traits between T1 and T2

#### Blood biomarkers

In the PR group, we report changes in nine out of the 16 blood biomarkers measured (Table 2, Fig. 1) including substantial reductions in total and LDL cholesterol, urea, creatinine, ALT, and γ-GT. Furthermore, we report a notable decrease in levels of CRP (72.8%, p = 0.001). A small decrease is also reported in levels of HDL cholesterol. Increased levels were only observed for ALP (Table 2). In NR individuals we report only small changes in levels of creatinine and γ-GT (Table 2, Fig. 1).

**Table 2.** Multivariable regression models exploring within-group changes in blood biomarkers between two dietary periods; omnivory and short-term APR (reference period). Between-group differences within each dietary period are also reported.

|  | *PR group* |  |  | *NR group* |  |  | *Between-group estimate (T1)* | *Between-group estimate (T2)* |
| --- | --- | --- | --- | --- | --- | --- | --- | --- |
| **Measured traits**<br>**Blood biomarkers** | **Within-group Estimate** | **P-value** | **95% CI** | **Within-group Estimate** | **P-value** | **95% CI** |  |  |
| Total cholesterol (mmol/L) | -0.3 | <0.0001*** | -0.4, -0.2 | 0.03 | 0.4 | -0.04, 0.1 | 0.04 | -0.3*** |
| LDL (mmol/L) | -0.3 | <0.0001*** | -0.4, -0.2 | -0.04 | 0.3 | -0.1, 0.03 | -0.001 | -0.3*** |
| HDL (mmol/L) | -0.07 | <0.0001*** | -0.1, -0.04 | -0.01 | 0.4 | -0.04, 0.01 | 0.05 | -8.8e-3 |
| Triglycerides (mmol/L) | 1.3e-3 | 0.04*† | 1.03e-4, 2.6e-3 | 5.2e-4 | 0.4 | -6.5e-4, 1.8e-3 | -1.3e-4 | 4.1e-4 |
| Glucose (mmol/L) | 2e-4 | 0.5 | -4e-4, 8e-3 | 6e-4 | 0.03*† | 1e-4, 2e-3 | -2.1e-4 | 0.001 |
| Insulin (mU/L) | 0.07 | 0.03*† | 0.01, 0.1 | 0.04 | 0.2 | -0.02, 0.1 | 0.008 | 0.04 |
| HbA1c (%) | -0.03 | 0.05*† | -0.06, -5e-4 | 0.002 | 0.9 | -0.03, 0.03 | -0.02 | 0.05 |
| Urea (mg/dL) | -5.8 | <0.0001*** | -6.9, -4.8 | 0.2 | 0.7 | -0.8, 1.2 | 0.7 | -4.9*** |
| Uric acid (mg/dL) | -0.06 | 0.2 | -0.1, 0.03 | -0.02 | 0.6 | -0.1, 0.06 | 0.1 | -0.1 |
| Creatinine (mg/dL) | -0.06 | <0.0001*** | -0.07, -0.04 | -0.02 | 0.01* | -0.04, -0.004 | -0.01 | -0.05** |
| AST (U/L) | 0.04 | 0.03*† | 4e-3, 0.07 | 0.02 | 0.3 | -0.02, 0.05 | 0.005 | -0.03 |
| ALT (U/L) | -0.09 | 0.0001*** | -0.1, -0.05 | -0.04 | 0.06 | -0.09, 0.002 | -0.01 | -0.07 |
| γ-GT (U/L) | -0.1 | <0.0001*** | -0.2, -0.1 | -0.09 | <0.0001*** | -0.1, -0.06 | -0.06 | -0.1*† |
| ALP (U/L) | 2.5 | <0.0001*** | 1.3, 3.7 | 0.5 | 0.4 | -0.6, 1.7 | 3.6*† | 5.9** |
| TSH (mIU/L) | 0.05 | 0.5 | -0.08, 0.2 | 0.1 | 0.04*† | 0.01, 0.3 | 0.3*† | -0.2 |
| CRP (mg/L) | -1.3 | 0.001** | -2.1, -0.5 | -0.4 | 0.3 | -1.3, 0.4 | 0.2 | -0.6 |
| **HOMA** |  |  |  |  |  |  |  |  |
| HOMA-B (--) | 13.3 | 0.02*† | 2.1, 24.5 | -3.8 | 0.5 | -14.9, 7.2 | -3.4 | 12.4 |
| HOMA-IR (--) | 0.2 | 0.4 | -0.3, 0.7 | -0.2 | 0.4 | -0.7, 0.3 | -0.5 | -0.01 |

*P*-value: \* ≤ 0.05, \*\* ≤ 0.001, \*\*\* ≤ 0.0001. Analyses adjusted for age, sex, BMI and medication use. † = not significant after Tukey's correction Abbreviations: LDL = Low-Density Lipoprotein, HDL = High-Density Lipoprotein, HbA1c = Hemoglobin A1c, AST = Aspartate Aminotransferase, ALT = Alanine Aminotransferase, γ-GT = Glutamyltransferase, ALP = Alkaline Phosphatase, TSH = Thyroid-Stimulating Hormone, CRP = C-Reactive Protein, HOMA = Homeostasis Model Assessment, HOMA-B =HOMA-Beta-Cell Function, HOMA-IR = HOMA-Insulin Resistance

**Figure 1:**
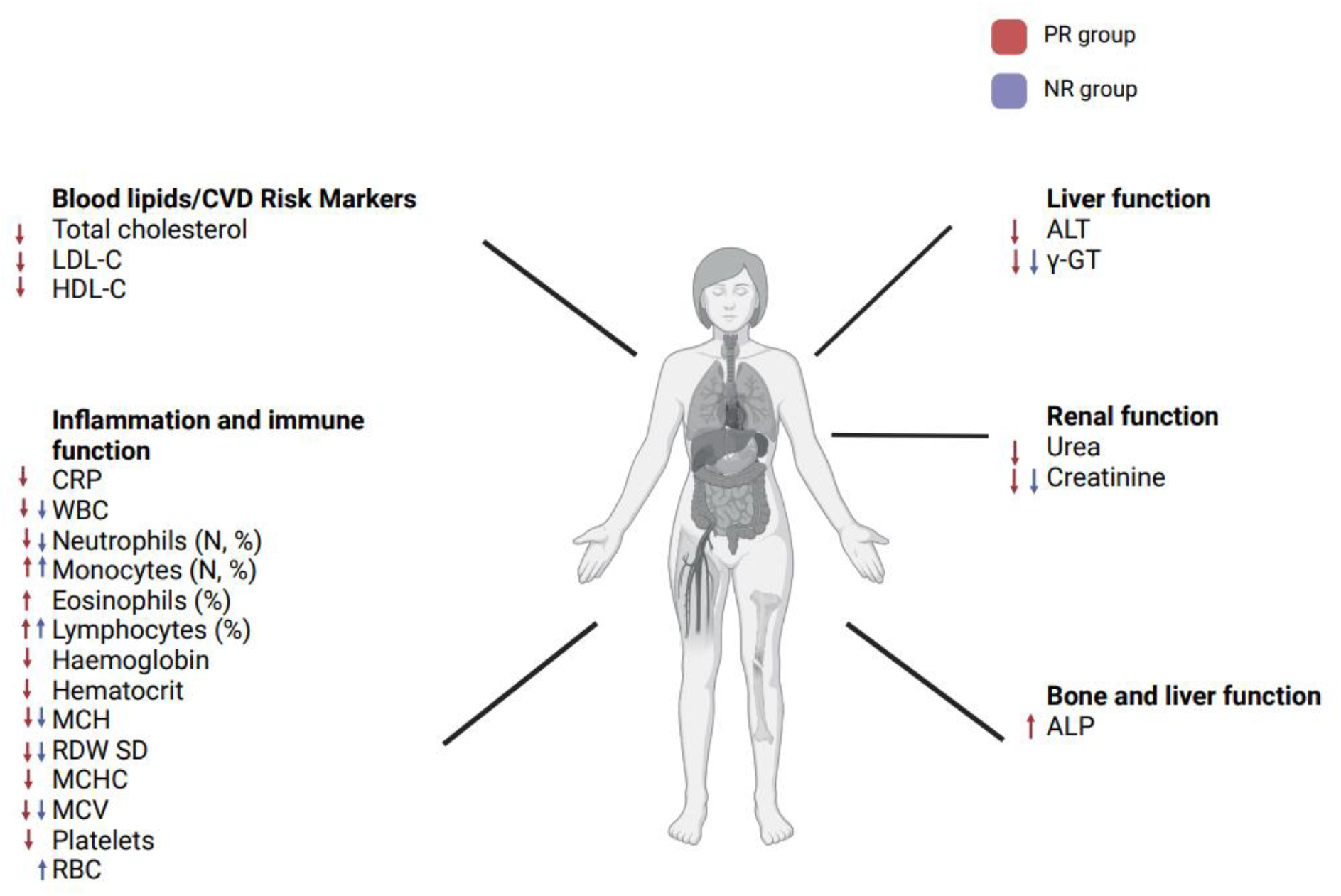
Changes in measured traits recorded from T1 to T2 for each dietary group. In PR individuals (red), short term-APR was associated with mostly beneficial changes in multiple markers of health, including prominent reductions in levels of total and LDL cholesterol and of CRP. Fewer changes, likely capturing seasonal effects on the immune system, were recorded for the NR group (blue). Abbreviations: PR, periodically-restricted; NR, non-restricted; LDL-C, Low-Density Lipoprotein-Cholesterol; HDL-C, High-Density Lipoprotein-Cholesterol; HbA1c = Hemoglobin A1c; HOMA-B, HOMA-Beta-Cell Function; CRP, C-Reactive Protein; WBC, White Blood Cells; MCH, Mean Corpuscular Hemoglobin; RDW CV, Red Cell Distribution Width Coefficient Of Variation; RDW SD, Red Cell Distribution Width Standard Deviation; MCHC, Mean Corpuscular Hemoglobin Concentration; MCV, Mean Corpuscular Volume; P-LCR, Platelet-Large Cell Ratio; AST, Aspartate Aminotransferase; ALT, Alanine Aminotransferase; γ-GT, Glutamyltransferase; ALP, Alkaline Phosphatase.

#### CBC traits

We report changes in 13 and 10 out of the 23 CBC traits measured, in the PR and NR groups respectively (Table 3, Fig.1) with both dietary groups displaying a reduction in total white blood (WBC) counts. Furthermore, both groups, display changes in lymphocyte %, monocyte %, neutrophils and neutrophil % (Table 3) while for red blood cell traits we report a decrease in MCH, MCV, and RDW SD (Table 3, Fig.1). In the PR group only, we report an increase in eosinophil %, and decreased levels of hemoglobin, haematocrit, MCHC, and platelets. In the NR group only, we report a small increase in monocytes and red blood cells (RBC) (Table 3, Fig.1).

**Table 3.**
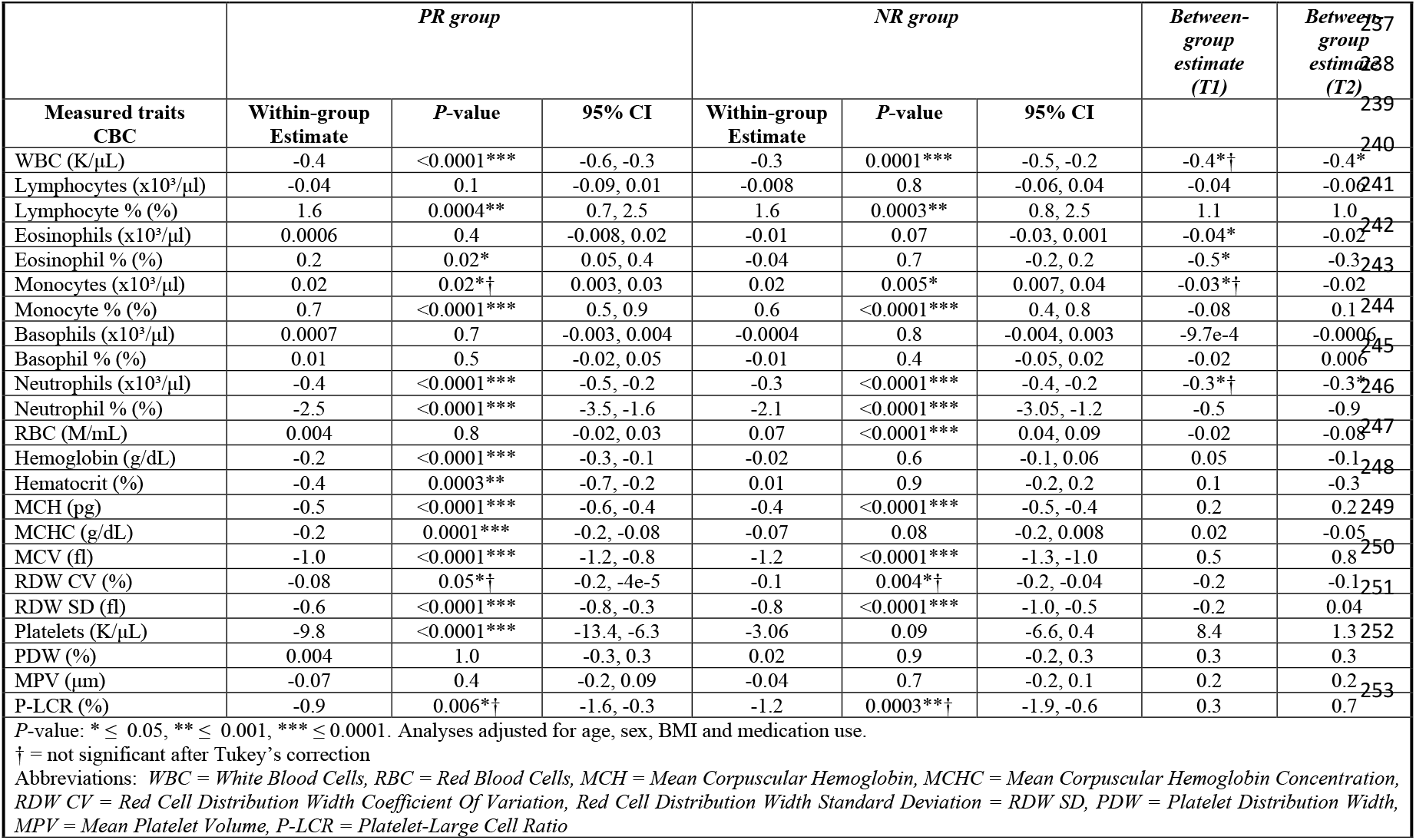
Multivariable regression models exploring within-group changes in CBC traits between two dietary periods; omnivory and short-term APR (reference period). Between-group differences within each dietary period are also reported.

| Measured traits<br>CBC | *PR group* |  |  | *NR group* |  |  | *Between-group*<br>*estimate*<br>*(T1)* | *Between-group*<br>*estimate*<br>*(T2)* |
| --- | --- | --- | --- | --- | --- | --- | --- | --- |
|  | Within-group<br>Estimate | *P*-value | 95% CI | Within-group<br>Estimate | *P*-value | 95% CI |  |  |
| WBC (K/μL) | -0.4 | <0.0001*** | -0.6, -0.3 | -0.3 | 0.0001*** | -0.5, -0.2 | -0.4*† | -0.4* |
| Lymphocytes (x10<sup>3</sup>/μl) | -0.04 | 0.1 | -0.09, 0.01 | -0.008 | 0.8 | -0.06, 0.04 | -0.04 | -0.06 |
| Lymphocyte % (%) | 1.6 | 0.0004** | 0.7, 2.5 | 1.6 | 0.0003** | 0.8, 2.5 | 1.1 | 1.0 |
| Eosinophils (x10<sup>3</sup>/μl) | 0.0006 | 0.4 | -0.008, 0.02 | -0.01 | 0.07 | -0.03, 0.001 | -0.04* | -0.02 |
| Eosinophil % (%) | 0.2 | 0.02* | 0.05, 0.4 | -0.04 | 0.7 | -0.2, 0.2 | -0.5* | -0.3 |
| Monocytes (x10<sup>3</sup>/μl) | 0.02 | 0.02*† | 0.003, 0.03 | 0.02 | 0.005* | 0.007, 0.04 | -0.03*† | -0.02 |
| Monocyte % (%) | 0.7 | <0.0001*** | 0.5, 0.9 | 0.6 | <0.0001*** | 0.4, 0.8 | -0.08 | 0.1 |
| Basophils (x10<sup>3</sup>/μl) | 0.0007 | 0.7 | -0.003, 0.004 | -0.0004 | 0.8 | -0.004, 0.003 | -9.7e-4 | -0.0006 |
| Basophil % (%) | 0.01 | 0.5 | -0.02, 0.05 | -0.01 | 0.4 | -0.05, 0.02 | -0.02 | 0.006 |
| Neutrophils (x10<sup>3</sup>/μl) | -0.4 | <0.0001*** | -0.5, -0.2 | -0.3 | <0.0001*** | -0.4, -0.2 | -0.3*† | -0.3* |
| Neutrophil % (%) | -2.5 | <0.0001*** | -3.5, -1.6 | -2.1 | <0.0001*** | -3.05, -1.2 | -0.5 | -0.9 |
| RBC (M/mL) | 0.004 | 0.8 | -0.02, 0.03 | 0.07 | <0.0001*** | 0.04, 0.09 | -0.02 | -0.08 |
| Hemoglobin (g/dL) | -0.2 | <0.0001*** | -0.3, -0.1 | -0.02 | 0.6 | -0.1, 0.06 | 0.05 | -0.1 |
| Hematocrit (%) | -0.4 | 0.0003** | -0.7, -0.2 | 0.01 | 0.9 | -0.2, 0.2 | 0.1 | -0.3 |
| MCH (pg) | -0.5 | <0.0001*** | -0.6, -0.4 | -0.4 | <0.0001*** | -0.5, -0.4 | 0.2 | 0.2 |
| MCHC (g/dL) | -0.2 | 0.0001*** | -0.2, -0.08 | -0.07 | 0.08 | -0.2, 0.008 | 0.02 | -0.05 |
| MCV (fl) | -1.0 | <0.0001*** | -1.2, -0.8 | -1.2 | <0.0001*** | -1.3, -1.0 | 0.5 | 0.8 |
| RDW CV (%) | -0.08 | 0.05*† | -0.2, -4e-5 | -0.1 | 0.004*† | -0.2, -0.04 | -0.2 | -0.1 |
| RDW SD (fl) | -0.6 | <0.0001*** | -0.8, -0.3 | -0.8 | <0.0001*** | -1.0, -0.5 | -0.2 | 0.04 |
| Platelets (K/μL) | -9.8 | <0.0001*** | -13.4, -6.3 | -3.06 | 0.09 | -6.6, 0.4 | 8.4 | 1.3 |
| PDW (%) | 0.004 | 1.0 | -0.3, 0.3 | 0.02 | 0.9 | -0.2, 0.3 | 0.3 | 0.3 |
| MPV (μm) | -0.07 | 0.4 | -0.2, 0.09 | -0.04 | 0.7 | -0.2, 0.1 | 0.2 | 0.2 |
| P-LCR (%) | -0.9 | 0.006*† | -1.6, -0.3 | -1.2 | 0.0003**† | -1.9, -0.6 | 0.3 | 0.7 |

*P*-value: \* $\leq 0.05$ , \*\* $\leq 0.001$ , \*\*\* $\leq 0.0001$ . Analyses adjusted for age, sex, BMI and medication use. † = not significant after Tukey's correction Abbreviations: WBC = White Blood Cells, RBC = Red Blood Cells, MCH = Mean Corpuscular Hemoglobin, MCHC = Mean Corpuscular Hemoglobin Concentration, RDW CV = Red Cell Distribution Width Coefficient Of Variation, Red Cell Distribution Width Standard Deviation = RDW SD, PDW = Platelet Distribution Width, MPV = Mean Platelet Volume, P-LCR = Platelet-Large Cell Ratio

#### Anthropometric traits and blood pressure

A very small decrease in BMI was observed in the PR group only, from T1 to T2. Additionally, a small decrease in DBP was observed for both dietary groups (Table 4).

**Table 4.** Multivariable regression models exploring within-group changes in anthropometric and blood pressure traits between two dietary periods; omnivory and short-term APR (reference period). Between-group differences within each dietary period are also reported.

|  | PR group |  |  | NR group |  |  | Between-group estimate (T1) | Between-group estimate (T2) |
| --- | --- | --- | --- | --- | --- | --- | --- | --- |
| Measured traits | Within-group Estimate | P-value | 95% CI | Within-group Estimate | P-value | 95% CI |  |  |
| Weight (kg) | -0.2 | 0.5 | -0.7, 0.4 | 1.0 | 0.0002**† | 0.5, 1.5 | 2.1 | 1.04 |
| BMI (kg/m²) | -0.02 | 0.5¥ | -0.07, 0.04 | 0.05 | 0.07 | -0.004, 0.002 | 0.2*† | 0.1 |
| SBP (mmHg) | -1.9 | 0.06 | -3.9, 0.09 | -2.2 | 0.03*† | -4.2, -0.2 | 0.4 | -0.8 |
| DBP (mmHg) | -2.5 | 0.0002** | -3.7, -1.2 | -2.2 | 0.0006** | -3.5, -1.0 | 0.02 | -1.04 |

P-value: \* ≤ 0.05, \*\* ≤ 0.001, \*\*\* ≤ 0.0001 † = not significant after Tukey’s correction ¥ = statistically significant after Tukey’s correction (β= -9.6e-16, p < 0.0001) Abbreviations: BMI = Body Mass Index; SBP = Systolic Blood Pressure; DBP = Diastolic Blood Pressure

#### 10-year CVD risk score

Upon APR, the CVD risk score for PR individuals (mean age = 56.0 years old) decreased from 5.3% to 4.9% (p = 0.02) resulting in a shift of PR individuals from the high to the low-to-moderate risk group, according to the latest ESC guidelines ^16^. No change in CVD risk score was detected for NR individuals (from 5.2% to 5.0%, p = 0.2).

#### Between-group differences of measured traits at T1 and T2

At T1, when both dietary groups were on an omnivorous diet, we report no differences in blood biomarker levels between PR and NR individuals (Table 2). At T2 (restriction timepoint), we report lower levels of total and LDL cholesterol, urea and creatinine for the PR group, but higher levels of ALP (Table 2). We also report differences for CBC traits with PR individuals displaying lower levels of eosinophils and eosinophil % at T1, and lower levels of WBC and neutrophils at T2 compared to the NR group (Table 3). No differences were observed in anthropometric or blood pressure traits between dietary groups.

## Discussion

### Principal findings

In the present study we find that short-term APR is linked to a substantial decrease in the levels of total and LDL cholesterol, to a 72.8% decrease in normal-range concentration of CRP, to improvements in markers of renal and liver function, and to a reduction of CVD risk (Fig. 2). Most changes were in a direction associated with positive effects on health, with the exception of ALP, a proxy of liver and bone health ^21^, which increased upon APR and may reflect negative effects of this dietary pattern on bone health. Given that this flexitarian type of diet is relatively easy to implement, further work is required to evaluate its long-term effects on human health, including its impact on liver fat accumulation ^22^, and its potential for lowering of risk for chronic diseases.

**Figure 2:**
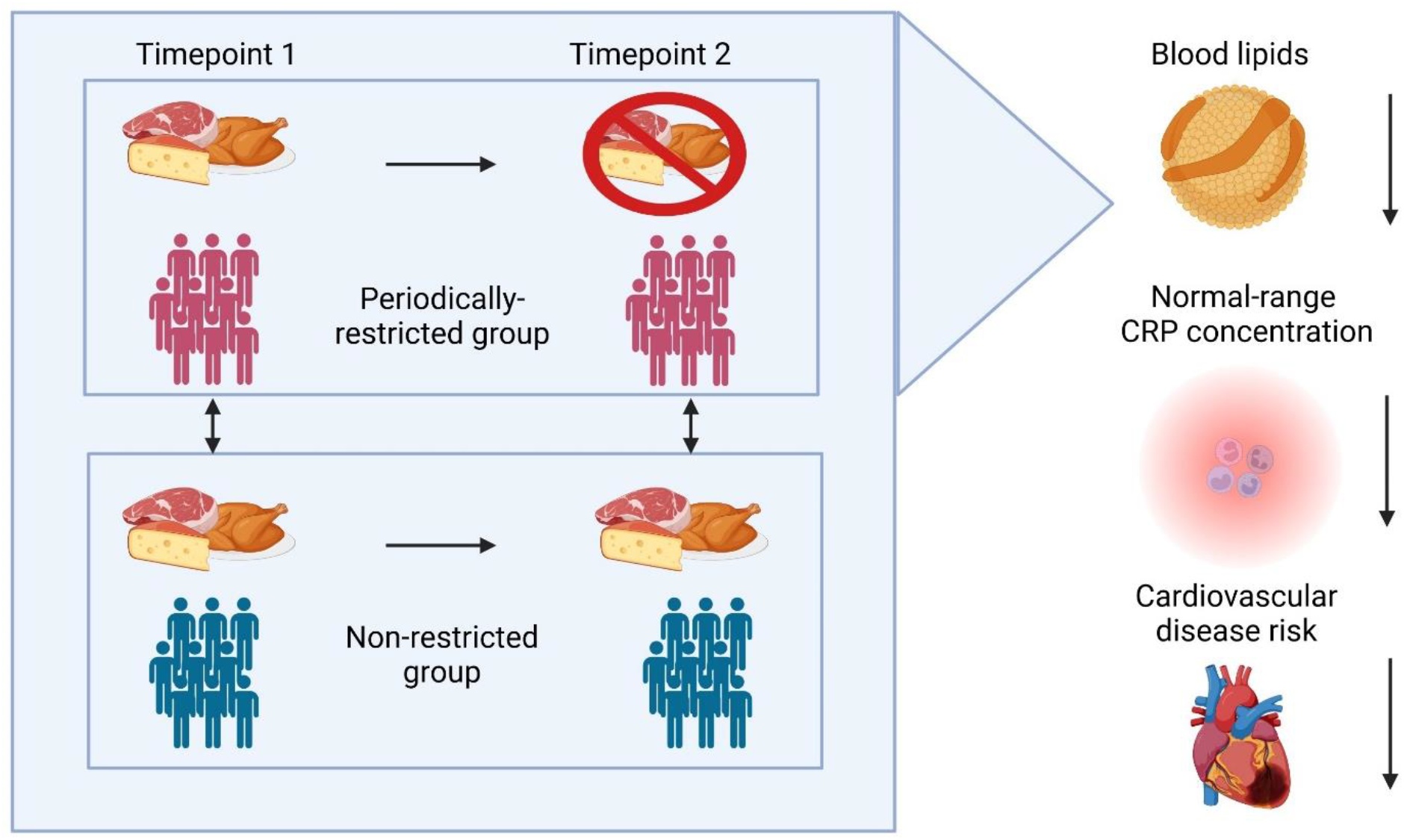
Effects of short-term APR on proxies of human health. A three-to-four week course of APR is associated with reduced levels of total and LDL cholesterol, reduced concentration of normal-range CRP and a reduction in the 10-year CVD risk.

### Blood lipids

We report rapid reductions of both total and LDL cholesterol levels upon APR, which drive a reduced risk for CVD risk following restriction, as measured by SCORE2. This reduction in CVD risk shifts PR individuals from a high to a low-to-moderate risk CVD category. SCORE2 ^20^, an index based on age, sex, smoking habits, SBP, total cholesterol and HDL cholesterol, is a widely accepted tool used by clinicians for the prediction of 10-year risk for fatal and non-fatal CVD events. In the present study we find that CVD risk scores changed from 5.3% to 4.9% for the PR group and from 5.2% to 5.0% for the NR group. Despite being only of slightly higher magnitude, the change in risk score was significant only for the PR group. Notably, this group is on average older and has a higher BMI (Table 1) compared to the NR group. Considering that aging is a non-modifiable risk factor for CVD events ^23^ and that increasing BMI is one of the leading risk factors for CVD ^24^, the documented shift of PR individuals to a lower CVD risk category upon short-term APR is of key importance as it demonstrates the potential of moderate lifestyle interventions to mitigate CVD risk.

Furthermore, while APR was linked to lower levels of total and LDL cholesterol, it was also linked to a slight decrease of HDL cholesterol levels. This effect on blood lipids has been reported previously ^9,25^ and may stem from the substantial reduction of animal fat intake. Given the impact of dietary intake on CVD risk, incorporating dietary information in CVD prediction models, in addition to the classical CVD risk factors, could improve robustness of patient classification and of CVD risk prediction.

Despite the relatively short duration of APR in the present study, the reduction in total and LDL cholesterol reported (0.3 mmol/L) was larger or comparable to changes reported in meta-analyses of RCTs studying similar types of dietary interventions, including vegan diets of three times longer duration ^6,7^. For example, in a meta-analysis of individuals at high risk of CVD, Rees et al. report reductions of 0.24 mmol/L and 0.22 mmol/L for total and LDL cholesterol respectively, after at least 12 weeks of a vegan diet ^7^. Similarly, Selinger et al ^9^ report a decrease of 0.48 mmol/L in LDL cholesterol in a meta-analysis of healthy individuals following a vegan diet for at least 12 weeks. Meta-analyses for similar dietary interventions in individuals with overweight, diabetes or at high risk for CVD report reductions in levels of total and LDL cholesterol ranging from 0.07 to 0.24 mmol/L ^6,9^. Through FastBio, we therefore unveil comparable effects on blood lipids through an intervention that is easy to apply and is of a shorter duration.

To put our finding in context, statin use lowers LDL cholesterol levels by an average of 1.8 mmol/L, reducing risk of ischemic heart disease (IHD) events by 60% and of stroke by 17% ^26^. Although the effects reported in the present study are of lower magnitude, they are nevertheless rapid and substantial suggesting that APR can be applied in combination with lipid-lowering therapy as a preventive measure against CVD.

### Markers of inflammation and immune system cells

Following APR, a 72.8% reduction was recorded for circulating, normal-range levels of CRP in PR individuals, suggesting a tempering effect of this diet on inflammation. Some evidence in the same direction is provided by meta-analysis of cross-sectional studies showing that permanent vegans have lower concentrations of CRP compared to omnivorous individuals ^27^. Importantly, the design of the present study enabled us to capture likely seasonal effects on the immune system. From autumn (T1) to spring (T2) both dietary groups undergo changes in total WBC counts, lymphocyte %, monocyte %, neutrophil counts and neutrophil %, in line with findings from the UK Biobank ^18^ (with the exception of monocyte %). In addition to seasonal findings, we report higher levels of eosinophil % and lower levels of hemoglobin, hematocrit, MCHC and platelets in the PR group only. The decrease observed in platelets, which in addition to their role in haemostasis and thrombosis can initiate or modulate protective or adverse immune functions ^28^, is in line with findings from a smaller interventional study on 53 individuals who consumed a vegan diet for four weeks ^12^. The same study reported that numbers of monocytes and neutrophils, but not lymphocytes, decreased upon animal product abstinence, with no changes reported for the dietary group following a meat-rich diet. Furthermore, a recent study on vegan versus ketogenic diets in humans ^13^ reported that neutrophils were the main driver for the upregulation of pathways linked to innate immunity, following a two-week vegan diet.

Overall, the reduction in CRP levels uncovered in the present study is of key importance as translational research studies and clinical trials highlight the central role of inflammation in the development of atherosclerosis ^29^. Although successful pharmacological approaches for the past 40 years were centred around LDL cholesterol lowering only, clinicians now have strong evidence that inhibition of inflammatory targets including CRP, IL-6 and IL-1β can protect against atherosclerosis ^30^. An example of such a trial is CANTOS, which targeted IL-1β through an antibody resulting in reduction of major CVD events by 15-17% ^30^. The inhibition of IL-1β was directly linked to the reduction in IL-6 and CRP. Therefore, elucidating the modulatory effect of dietary interventions on the immune system remains a largely untapped resource of enormous potential for the prevention and treatment of disease.

### Renal, liver and bone health

Building on knowledge from cross-sectional studies ^5^, we report rapid reductions in levels of urea and creatinine associated with APR, likely reflecting decreased protein intake. We also observed a decrease in levels of ALT upon restriction, suggesting lower levels of fat deposition in the liver ^31,32^ and potentially protective effects against hepatic steatosis ^33^. Furthermore, at T2, levels of γ-GT are reduced for both PR and NR groups, likely reflecting lower levels of alcohol consumption. Although this was expected for the PR group, lower γ-GT levels in the NR group may be a consequence of awareness bias following participation in the study at T1, and subsequent curbing of alcohol consumption. Notably, ALP levels increased upon APR suggesting possible negative effects on bone or liver health ^21^. Large cross-sectional cohort studies have shown that permanent vegans have a higher risk of bone fractures ^11^. We suggest that higher ALP levels may reflect negative effects of periodic APR on bone health, but further studies are needed to establish the magnitude of effect of this dietary pattern on bone health by using more representative tests such as quantification of bone mineral density (BMD), bone-specific ALP, C-terminal telopeptide (CTx) and parathyroid hormone ^34^.

### Strengths and limitations

The key strength of the FastBio study is that it comprises a unique group of individuals who have followed a specific dietary pattern, in a consistent manner, for at least a decade. FastBio PR individuals therefore are highly homogeneous in the foodstuffs they abstain from, and we hypothesize that restriction from these products drives a large fraction of reported effects. Despite the enormous potential of dietary interventions to contribute to the prevention and treatment of disease, attaining this level of homogeneity in dietary studies is challenging and adhering consistently to specific diets for prolonged periods of time can prove difficult, resulting in substantial heterogeneity in the dietary groups studied. Furthermore, in contrast to studies reporting on a few measurements, FastBio is one of the most comprehensive studies addressing the effects of APR on markers of health, reporting on 16 biomarker and 23 CBC traits, as well as on anthropometric and blood pressure traits. Importantly, the FastBio study enables us to disentangle effects driven by seasonality and given the sampling strategy applied, to minimise circadian effects on traits such as WBCs and CRP that are known to display significant daily variation ^18^.

A limitation of the FastBio study is that traits were measured at two distinct timepoints (approximately five months apart), but not immediately prior to initiation of restriction, meaning that we do not have typical baseline measurements. Prior to initiation of restriction however, PR individuals had undergone a period of omnivory that was similar in length to the period of omnivory prior to T1. This supports the idea that prior to restriction at T2, for the traits measured in the present study, the PR group had a similar biological profile to that measured at T1. Furthermore, most blood biomarkers measured are known to capture relatively acute dietary effects, meaning that with the present measurements we are underpowered to detect likely long-term effects of periodic restriction of animal products on markers of human health. Another limitation of our study is that we report on levels of inflammation through quantification of CRP levels instead of high-sensitivity CRP (hs-CRP), which is more commonly used to detect the risk for inflammatory conditions ^35^. Finally, although we have adjusted for confounding factors to the best of our ability, some residual effects may persist in reported results.

## Conclusions

Over the past decades flexible dietary patterns that are compatible with food variety, cultural traditions, and personal preferences are growing in popularity ^36^. The combination of these elements can be found in flexitarian type of diets that are less restrictive and have a likely positive impact on human health^15^. The dietary pattern presented here constitutes such an approach and results in a biomarker profile compatible with positive effects on health and a reduction of CVD risk. Although further work is necessary to address potential negative effects on bone health and to evaluate long-term effects, harnessing flexitarian type of dietary interventions is a promising approach that can be applied for disease prevention, and that can inform on future public health dietary guidelines.

## Data availability

The data underlying this article will be shared on reasonable request to the corresponding author.

## Abbreviations

PR: Periodically restricted
NR: Non-restricted
T1: Timepoint 1
T2: Timepoint 2
BMI: Body Mass Index
SBP: Systolic Blood Pressure
DBP: Diastolic Blood Pressure
LDL: Low-Density Lipoprotein
HDL: High-Density Lipoprotein
Hba1c: Hemoglobin A1c
AST: Aspartate Aminotransferase
ALT: Alanine Aminotransferase
γ-GT: Glutamyltransferase
ALP: Alkaline Phosphatase
TSH: Thyroid-Stimulating Hormone
CRP: C-Reactive Protein
HOMA: Homeostasis Model Assessment
HOMA-B: HOMA-Beta-Cell Function
HOMA-IR: HOMA-Insulin Resistance
WBC: White Blood Cells
RBC: Red Blood Cells
MCH: Mean Corpuscular Hemoglobin
MCHC: Mean Corpuscular Hemoglobin Concentration
RDW CV: Red Cell Distribution Width Coefficient of Variation
RDW: SD Red Cell Distribution Width Standard Deviation
PDW: Platelet Distribution Width
MPV: Mean Platelet Volume
P-LCR: Platelet-Large Cell Ratio
MAF: Minor Allele Frequency
HWE: Hardy-Weinberg Equilibrium
CVD: Cardiovascular disease

## Author contributions

EML: Conceptualization, Methodology, Formal analysis, Writing – original draft. AS: Formal analysis, Writing – Review & Editing. PB: Formal analysis, Writing – Review & Editing. SG: Data acquisition, Material Preparation, Writing – Review & Editing. AD: Data acquisition, Material Preparation, Writing – Review & Editing. MA: Data acquisition, Material Preparation, Writing – Review & Editing. PR: Interpretation, Writing – Review & Editing. IK: Conceptualization, Methodology, Interpretation, Writing – Review & Editing. ND: Conceptualization, Methodology, Interpretation, Writing – Review & Editing. NS: Conceptualization, Methodology, Interpretation, Writing – Review & Editing. MY: Interpretation, Writing – Review & Editing. KR: Data acquisition, Material Preparation, Writing – Review & Editing. ASD: Supervision, Funding acquisition, Conceptualization, Methodology, Data acquisition, Material Preparation, Writing – original draft.

## Acknowledgements

The authors would like to thank the FastBio study participants and the Interbalkan Hospital staff. We would also like to thank Dr Dimitrios Rouskas and Dr Loukas Kipouros for their invaluable help.

## REFERENCES

1 Jiang, Y. et al. Therapeutic Implications of Diet in Inflammatory Bowel Disease and Related Immune-Mediated Inflammatory Diseases. Nutrients 13, doi:10.3390/nu13030890 (2021).

2 Zhang, S. et al. Adherence to the EAT-Lancet diet, genetic susceptibility, and risk of type 2 diabetes in Swedish adults. Metabolism 141, 155401, doi:10.1016/j.metabol.2023.155401 (2023).

3 Dinu, M., Abbate, R., Gensini, G. F., Casini, A. & Sofi, F. Vegetarian, vegan diets and multiple health outcomes: A systematic review with meta-analysis of observational studies. Crit Rev Food Sci Nutr 57, 3640–3649, doi:10.1080/10408398.2016.1138447 (2017).

4 Medawar, E., Huhn, S., Villringer, A. & Veronica Witte, A. The effects of plant-based diets on the body and the brain: a systematic review. Transl Psychiatry 9, 226, doi:10.1038/s41398-019-0552-0 (2019).

5 Tong, T. Y. N., Perez-Cornago, A., Bradbury, K. E. & Key, T. J. Biomarker Concentrations in White and British Indian Vegetarians and Nonvegetarians in the UK Biobank. J Nutr 151, 3168–3179, doi:10.1093/jn/nxab192 (2021).

6 Termannsen, A. D. et al. Effects of vegan diets on cardiometabolic health: A systematic review and meta-analysis of randomized controlled trials. Obes Rev 23, e13462, doi:10.1111/obr.13462 (2022).

7 Rees, K., Al-Khudairy, L., Takeda, A. & Stranges, S. Vegan dietary pattern for the primary and secondary prevention of cardiovascular diseases. Cochrane Database Syst Rev 2, Cd013501, doi:10.1002/14651858.CD013501.pub2 (2021).

8 Yokoyama, Y., Levin, S. M. & Barnard, N. D. Association between plant-based diets and plasma lipids: a systematic review and meta-analysis. Nutr Rev 75, 683–698, doi:10.1093/nutrit/nux030 (2017).

9 Selinger, E. et al. Evidence of a vegan diet for health benefits and risks – an umbrella review of meta-analyses of observational and clinical studies. Critical Reviews in Food Science and Nutrition, 1–11, doi:10.1080/10408398.2022.2075311 (2022).

10 Key, T. J., Papier, K. & Tong, T. Y. N. Plant-based diets and long-term health: findings from the EPIC-Oxford study. Proc Nutr Soc 81, 190–198, doi:10.1017/s0029665121003748 (2022).

11 Tong, T. Y. N. et al. Vegetarian and vegan diets and risks of total and site-specific fractures: results from the prospective EPIC-Oxford study. BMC Med 18, 353, doi:10.1186/s12916-020-01815-3 (2020).

12 Lederer, A. K. et al. Vegan diet reduces neutrophils, monocytes and platelets related to branched-chain amino acids - A randomized, controlled trial. Clin Nutr 39, 3241–3250, doi:10.1016/j.clnu.2020.02.011 (2020).

13 Link, V. M. et al. Differential peripheral immune signatures elicited by vegan versus ketogenic diets in humans. Nat Med 30, 560–572, doi:10.1038/s41591-023-02761-2 (2024).

14 Kjeldsen-Kragh, J. et al. Controlled trial of fasting and one-year vegetarian diet in rheumatoid arthritis. Lancet (London, England) 338, 899–902, doi:10.1016/0140-6736(91)91770-u (1991).

15 Derbyshire, E. J. Flexitarian Diets and Health: A Review of the Evidence-Based Literature. Front Nutr 3, 55, doi:10.3389/fnut.2016.00055 (2016).

16 Visseren, F. L. J. et al. 2021 ESC Guidelines on cardiovascular disease prevention in clinical practice: Developed by the Task Force for cardiovascular disease prevention in clinical practice with representatives of the European Society of Cardiology and 12 medical societies With the special contribution of the European Association of Preventive Cardiology (EAPC). Rev Esp Cardiol (Engl Ed) 75, 429, doi:10.1016/j.rec.2022.04.003 (2022).

17 Sarri, K. O., Linardakis, M. K., Bervanaki, F. N., Tzanakis, N. E. & Kafatos, A. G. Greek Orthodox fasting rituals: a hidden characteristic of the Mediterranean diet of Crete. Br J Nutr 92, 277–284, doi:10.1079/BJN20041197 (2004).

18 Wyse, C., O’Malley, G., Coogan, A. N., McConkey, S. & Smith, D. J. Seasonal and daytime variation in multiple immune parameters in humans: Evidence from 329,261 participants of the UK Biobank cohort. iScience 24, 102255, doi:10.1016/j.isci.2021.102255 (2021).

19 Primatesta, P., Falaschetti, E., Gupta, S., Marmot, M. G. & Poulter, N. R. Association between smoking and blood pressure: evidence from the health survey for England. Hypertension 37, 187–193, doi:10.1161/01.hyp.37.2.187 (2001).

20 group, S. w. & collaboration, E. S. C. C. r. SCORE2 risk prediction algorithms: new models to estimate 10-year risk of cardiovascular disease in Europe. European heart journal 42, 2439–2454, doi:10.1093/eurheartj/ehab309 (2021).

21 Kuo, T. R. & Chen, C. H. Bone biomarker for the clinical assessment of osteoporosis: recent developments and future perspectives. Biomark Res 5, 18, doi:10.1186/s40364-017-0097-4 (2017).

22 Georgakouli, K. et al. The Effects of Greek Orthodox Christian Fasting during Holy Week on Body Composition and Cardiometabolic Parameters in Overweight Adults. Diseases 10, doi:10.3390/diseases10040120 (2022).

23 Hamczyk, M. R., Nevado, R. M., Barettino, A., Fuster, V. & Andres, V. Biological Versus Chronological Aging: JACC Focus Seminar. J Am Coll Cardiol 75, 919–930, doi:10.1016/j.jacc.2019.11.062 (2020).

24 Global Cardiovascular Risk, C. et al. Global Effect of Modifiable Risk Factors on Cardiovascular Disease and Mortality. The New England journal of medicine 389, 1273–1285, doi:10.1056/NEJMoa2206916 (2023).

25 Kent, L. et al. The effect of a low-fat, plant-based lifestyle intervention (CHIP) on serum HDL levels and the implications for metabolic syndrome status - a cohort study. Nutr Metab (Lond) 10, 58, doi:10.1186/1743-7075-10-58 (2013).

26 Law, M. R., Wald, N. J. & Rudnicka, A. R. Quantifying effect of statins on low density lipoprotein cholesterol, ischaemic heart disease, and stroke: systematic review and meta-analysis. BMJ (Clinical research ed.) 326, 1423, doi:10.1136/bmj.326.7404.1423 (2003).

27 Menzel, J. et al. Systematic review and meta-analysis of the associations of vegan and vegetarian diets with inflammatory biomarkers. Sci Rep 10, 21736, doi:10.1038/s41598-020-78426-8 (2020).

28 Chen, Y., Zhong, H., Zhao, Y., Luo, X. & Gao, W. Role of platelet biomarkers in inflammatory response. Biomark Res 8, 28, doi:10.1186/s40364-020-00207-2 (2020).

29 Awan, Z. & Genest, J. Inflammation modulation and cardiovascular disease prevention. Eur J Prev Cardiol 22, 719–733, doi:10.1177/2047487314529350 (2015).

30 Ridker, P. M. From RESCUE to ZEUS: will interleukin-6 inhibition with ziltivekimab prove effective for cardiovascular event reduction? Cardiovasc Res 117, e138–e140, doi:10.1093/cvr/cvab231 (2021).

31 Despres, J. P. et al. Loss of abdominal fat and metabolic response to exercise training in obese women. Am J Physiol 261, E159–167, doi:10.1152/ajpendo.1991.261.2.E159 (1991).

32 Straznicky, N. E. et al. The effects of dietary weight loss with or without exercise training on liver enzymes in obese metabolic syndrome subjects. Diabetes Obes Metab 14, 139–148, doi:10.1111/j.1463-1326.2011.01497.x (2012).

33 Sanyal, A. J. et al. Pioglitazone, vitamin E, or placebo for nonalcoholic steatohepatitis. The New England journal of medicine 362, 1675–1685, doi:10.1056/NEJMoa0907929 (2010).

34 Greenblatt, M. B., Tsai, J. N. & Wein, M. N. Bone Turnover Markers in the Diagnosis and Monitoring of Metabolic Bone Disease. Clin Chem 63, 464–474, doi:10.1373/clinchem.2016.259085 (2017).

35 Lee, H. S. & Lee, J. H. Early elevation of high-sensitivity C-reactive protein as a predictor for cardiovascular disease incidence and all-cause mortality: a landmark analysis. Sci Rep 13, 14118, doi:10.1038/s41598-023-41081-w (2023).

36 Willett, W. et al. Food in the Anthropocene: the EAT-Lancet Commission on healthy diets from sustainable food systems. Lancet (London, England) 393, 447–492, doi:10.1016/S0140-6736(18)31788-4 (2019).

