## Supplementary Material for "Effects of short-term restriction of animal products on blood biomarkers and cardiovascular disease risk"

*Corresponding author:

### SUPPLEMENTARY TEXT

#### Text S1. Inclusion and exclusion criteria for FastBio study participants

Inclusion criteria:

- Healthy female and male subjects 18 – 75 years of age at the time of enrollment. One participant turned 76 between time of enrollment and first sampling timepoint (T1).
- Able to provide signed and dated informed consent. Willing to provide blood samples.
- Individuals who have followed periodic animal product restriction (APR) for at least ten years (for the PR group).
- Individuals who do not follow any kind of specific diet including veganism, vegetarianism, caloric restriction, intermittent fasting etc. (for the NR group).

Exclusion criteria:

- Use of antibiotic, antifungal, antiviral or antiparasitic drugs six months prior to T1.
- Acute disease, defined as the presence of a moderate or severe illness with or without fever, at sampling timepoints.
- Alcohol or drug abuse two years prior to T1.
- Participants who normally practice periodic restriction from animal products, but had to alter their diet for a specific reason (e.g. pregnancy).

#### Text S2. Dietary pattern of periodic APR as specified by the Greek Orthodox Church

Individuals following the dietary regimen of the Greek Orthodox Church practice restriction of animal products through abstinence from meat, fish, dairy products and eggs for 180-200 days annually. Consumption of shellfish and molluscs is permitted on all days of restriction. Restriction is practiced on Wednesdays and Fridays throughout the year (excluding the week immediately after Christmas, Easter and the Pentecost) and over four extended periods annually, as follows (Supplementary Figure S1):

- 40 days before Christmas (abstinence from meat, dairy product and eggs, consumption of fish is permitted except on Wednesdays and Fridays).
- 48 days before Easter (Lent) (abstinence from meat, fish, dairy products and eggs, consumption of fish is permitted only on March $25^{th}$ and on Palm Sunday).
- 0-30 days in June (abstinence from meat, dairy products and eggs, consumption of fish is permitted).

- 15 days before August 15th (Assumption) (abstinence from meat, fish, dairy products and eggs, consumption of fish is permitted only on August $6^{th}$).

**Text S3. Self-reported use of dietary supplements**

Self-reported use of dietary supplements for FastBio participants was as follows:
PR group: Mg (1 participant), Fe (2 participants), Vitamin B (1 participant) Calcium (3 participants), Vitamin B and Calcium (1 participant), Vitamin D (1 participant), amino acid supplement (1 participant)
NR group: Calcium (1 participant), Vitamin D (1 participant).

**Text S4. HOMA indices**

Prior to calculating HOMA indices, we inspected our data for glucose levels < 2.5mmol/L. No individuals were excluded based on this threshold. We calculated HOMA indices as follows [1, 2]:

$$\text{(a) } HOMA - B = (360 * Insulin)/(Glucose - 63)$$

$$\text{(b) } HOMA - IR = (Insulin * Glucose)/405$$

**Text S5: Missing data handling**

We excluded eight PR and 13 NR individuals from the complete sample (N = 411) because they were not present at T2. Findings are therefore from 200 PR and 211 NR individuals at T1 and 192 PR and 198 NR individuals at T2 (paired analysis: N = 390).

**Text S6. BMI and age categories**

Participants were grouped into four age categories based on representative classifications in the literature [3-5]:19-40, 41-50, 51-60 and 61-76 years old (Supplementary Table S2) and four BMI categories as indicated by the World Health Organization (WHO) [6]: < 18.5kg/$m^2$ underweight; 18.5 kg/$m^2$ ≤ BMI < 25 kg/$m^2$ normal weight; 25 kg/$m^2$ ≤ BMI < 30 kg/$m^2$, overweight; and ≥ 30 kg/$m^2$ obese (Supplementary Table S2).

**Text S7. Model adjustments**

Medications accounted for include drugs for blood lipids, blood pressure, blood glucose levels, osteoporosis, thyroid function, and anticoagulants. All models were run with: (a) medication use as a single, binary covariate (Yes/No), and (b) each medication type as a separate covariate with no differences in overall findings. We report estimates and CIs from the (b) approach.

In addition to the model adjustments described in the main text, we also adjusted for smoking and alcohol consumption with no impact on results. The results presented are therefore based on the models without these two adjustments. Self-reported levels of physical activity, which did not differ substantially between dietary groups in each timepoint and for each dietary group within timepoints, were not included in the models.

### SUPPLEMENTARY FIGURES

**Figure S1.** Indicative calendar showing days of APR (in gray) for PR individuals and timepoints of profiling for both dietary groups.

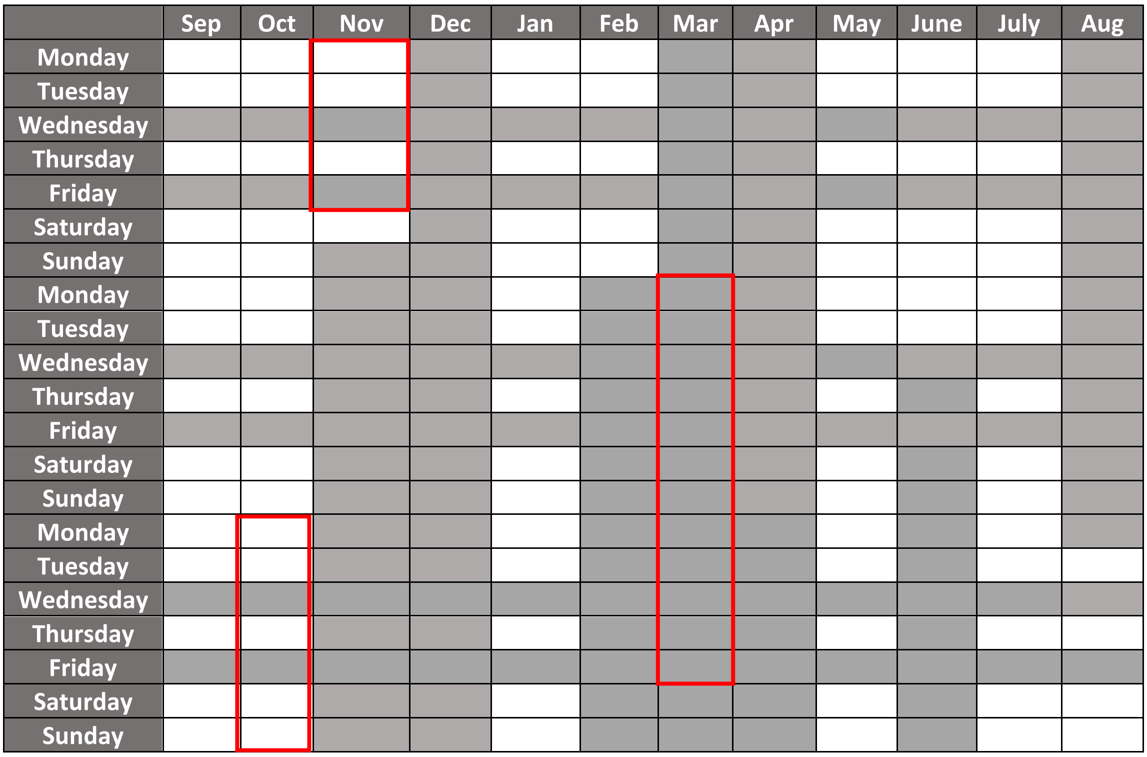

APR is practiced for 180-200 days annually, over four extended periods of restriction throughout the year as well as restriction on Wednesdays and Fridays of each week. When not practicing restriction, PR individuals follow an omnivorous diet. The time periods shown in red boxes are indicative of timepoint 1 (red boxes on left) and timepoint 2 (red box on right) of the FastBio study.

### SUPPLEMENTARY TABLES

**Table S1.** Blood biomarkers and CBC traits measured in the PR and NR dietary groups, at two timepoints.

| Measured traits | Mean ± sd | | | | Units | Normal range‡ |
|---|---|---|---|---|---|---|
| **Blood biomarkers** | T1 | | T2 | | | |
| | $PV_{(n=200)}$ | $NV_{(n=211)}$ | $PV_{(n=192)}$ | $NV_{(n=198)}$ | | |
| Total cholesterol | 5.1±0.9 | 4.9±0.8 | 4.7±0.9 | 5.0±0.9 | mmol/l | < 5.2 |
| LDL | 3.4±0.8 | 3.3±0.8 | 3.1±0.8 | 3.3±0.9 | mmol/l | < 3.4 |
| HDL | 1.6±0.4 | 1.6±0.4 | 1.5±0.4 | 1.5±0.4 | mmol/l | 0.9-1.6 |
| Triglycerides | 2.7±1.3 | 2.5±1.4 | 2.8±1.3 | 2.6±1.2 | mmol/l | < 3.9 |
| Glucose | 2.5±0.3 | 2.4±0.3 | 2.5±0.3 | 2.5±0.3 | mmol/l | 1.9-3.0 |
| Insulin | 9.8±9.0 | 9.4±13.4 | 10.4±11.0 | 9.4±9.3 | mU/L | 3.0-25.0 |
| HbA1c | 5.3±0.5 | 5.2±0.5 | 5.3±0.4 | 5.2±0.5 | % | 4.5-6.2 |
| Urea | 32±9.1 | 30±7.8 | 26±7.3 | 30±7.4 | mg/dl | 10-50 |
| Uric acid | 5.1±1.3 | 4.8±1.3 | 5.0±1.2 | 4.8±1.3 | mg/dl | 2.4-5.7 |
| Creatinine | 0.8±0.2 | 0.8±0.2 | 0.7±0.2 | 0.8±0.1 | mg/dl | 0.5-0.9 |
| AST | 19.1±7.7 | 18.5±6.0 | 20.0±7.0 | 18.7±4.9 | U/I | 10-32 |
| ALT | 20.8±12.5 | 20.5±12.2 | 19.1±11.5 | 19.4±9.0 | U/I | 10-33 |
| γ-GT | 19.3±15.9 | 18.8±17.3 | 16.1±11.0 | 17.4±15.9 | U/I | 5-36 |
| ALP | 66.0±17.3 | 60.0±16.4 | 68.0±17.0 | 60.0±15.8 | U/I | 35-104 |
| TSH | 2.0±1.3 | 1.8±1.0 | 2.0±1.1 | 1.9±1.1 | mIU/L | 0.4-4.9 |
| CRP | 2.6±2.7 | 2.2±2.4 | 2.2±2.3 | 2.4±3.6 | mg/L | 0-5 |
| **HOMA** | | | | | | |
| HOMA-B | 107.6±71.8 | 106.6±88.9 | 117.3±112.9 | 104.3±85.5 | --- | --- |
| HOMA-IR | 2.4±2.6 | 2.4±4.7 | 2.5±2.9 | 2.3±2.4 | --- | --- |
| **CBC** | | | | | | |
| WBC | 6.1±1.5 | 6.4±1.7 | 5.7±1.4 | 6.1±1.6 | K/µl | 4.0-11.0 |
| Lymphocytes | 2.0±0.6 | 2.0±0.6 | 1.9±0.6 | 2.0±0.6 | x10³/µl | 1.5-4.0 |
| Lymphocyte % | 33.0±6.5 | 32.0±7.1 | 34.0±7.1 | 33.0±7.5 | % | 20-40 |
| Eosinophils | 0.2±0.1 | 0.2±0.2 | 0.2±0.1 | 0.2±0.1 | x10³/µl | < 0.4 |
| Eosonophil % | 2.5±1.6 | 3.1±2.3 | 2.8±2.1 | 3.1±2.1 | % | 1 - 6 |
| Monocytes | 0.4±0.1 | 0.4±0.1 | 0.4±0.1 | 0.4±0.1 | x10³/µl | 0.1-0.9 |
| Monocyte % | 6.7±1.7 | 6.8±1.7 | 7.5±1.7 | 7.3±1.6 | % | 2-10 |
| Basophils | 0.003±0.02 | 0.003±0.02 | 0.003±0.02 | 0.003±0.02 | x10³/µl | < 0.1 |
| Basophil % | 0.03±0.2 | 0.04±0.2 | 0.04±0.2 | 0.03±0.2 | % | 0.3 - 1.0 |
| Neutrophils | 3.6±1.1 | 3.8±1.3 | 3.2±1.0 | 3.5±1.2 | x10³/µl | 2.0-7.7 |
| Neutrophil % | 58.1±6.9 | 58.5±7.7 | 55.5±7.7 | 56.3±8.1 | % | 40 - 75 |
| RBC | 4.9±0.5 | 4.9±0.5 | 4.9±0.5 | 5.0±0.5 | M/ml | 3.8-5.8 |
| Hemoglobin | 14.2±1.4 | 14.2±1.5 | 14.0±1.3 | 14.1±1.4 | g/dl | 11.5-16.5 |
| Hematocrit | 42.6±3.5 | 42.3±3.6 | 42.2±3.4 | 42.4±3.5 | % | 37-47 |
| MCH | 29.3±2.4 | 29.1±3.0 | 28.8±2.4 | 28.6±3.0 | Pg | 26.0 - 32.0 |
| MCHC | 33.4±0.9 | 33.4±1.0 | 33.2±1.0 | 33.3±1.0 | g/dL | 30-36 |
| MCV | 87.8±6.4 | 87.0±7.9 | 86.8±6.4 | 85.7±7.8 | Fl | 79.0 - 98.0 |
| RDW CV | 13.6±0.9 | 13.6±1.2 | 13.5±1.0 | 13.6±1.2 | % | 11.0 - 16.0 |
| RDW SD | 43.0±3.0 | 42.8±3.4 | 42.5±3.2 | 42.1±3.3 | Fl | 38-43 |
| Platelets | 246.0±51.1 | 236.8±48.3 | 236.3±50.0 | 235.1±50.0 | K/µl | 150.0-400.0 |
| PDW* | 12.7±2.8 | 12.4±3.7 | 12.9±1.9 | 13.0±1.9 | % | 9.0-17.0 |
| MPV* | 10.5±1.9 | 10.2±2.8 | 10.6±0.9 | 10.7±0.8 | Mm | 6-11 |
| P-LCR* | 30.6±8.8 | 30.2±10.4 | 30.2±7.3 | 30.5±6.9 | % | 13.0 - 43.0 |

‡ = IBH (Interbalkan Hospital) blood biomarker normal range, * = Paired data available for 190 PR and 193 NR individuals

*LDL = Low-Density Lipoprotein, HDL = High-Density Lipoprotein, HbA1c = Hemoglobin A1c, AST = Aspartate Aminotransferase, ALT = Alanine Aminotransferase, γ-GT = Glutamyltransferase, ALP = Alkaline Phosphatase, TSH = Thyroid-Stimulating Hormone, CRP = C-Reactive Protein, HOMA = Homeostasis Model Assessment, HOMA-B =HOMA- Beta-Cell Function , HOMA-IR = HOMA- Insulin Resistance; HOMA = Homeostasis Model*

*Assessment; WBC = White Blood Cells, RBC = Red Blood Cells, MCH = Mean Corpuscular Hemoglobin, MCHC = Mean Corpuscular Hemoglobin Concentration, MCV = Mean Corpuscular Volume, RDW CV = Red Cell Distribution Width Coefficient Of Variation, Red Cell Distribution Width Standard Deviation = RDW SD, PDW = Platelet Distribution Width, MPV = Mean Platelet Volume, P-LCR = Platelet-Large Cell Ratio*

**Table S2.** Study population characteristics at timepoint 1 (T1) and timepoint 2 (T2)[a].

| **Timepoint 1** | **Subgroups** | **PR (n = 200)** | **NR (n = 211)** | **Total (n = 411)** |
|---|---|---|---|---|
| **Age (years)** | | 51.5±13.5 | 45±13.1 | 48.1±13.6 |
| | Group 1: 19-40 (%) | 35 (17.5) | 82 (38.9) | 28.5 % |
| | Group 2: 41-50 (%) | 48 (24) | 43 (20.4) | 22.1 % |
| | Group 3: 51-60 (%) | 52 (26) | 55 (26) | 26% |
| | Group 4: 61-76 (%) | 65 (32.5) | 31 (14.7) | 23.4% |
| **Sex (%)** | Female | 108 (54) | 116 (55) | 54.5% |
| | Male | 92 (46) | 95 (45) | 45.5% |
| **Weight (kg)** | --- | 80±15.7 | 76±15.8 | 78±15.8 |
| **BMI (kg/m²)** | | 28.4±4.6 | 26.2±4.4 | 27.3±4.6 |
| | Group 1: < 18.5 (%) | 1 (0.5) | 6 (2.8) | 1.7% |
| | Group 2: 18.5-25 (%) | 39 (19.5) | 77 (36.5) | 28.2% |
| | Group 3: 25-30 (%) | 99 (49.5) | 89 (42.2) | 45.7% |
| | Group 4: ≥ 30 (%) | 61 (30.5) | 39 (18.5) | 24.3% |
| **Smoking (%)** | Non-smoker (%) | 187 (93.5) | 141 (66.8) | 79.8 % |
| | Smoker (%) | 13 (6.5) | 70 (33.2) | 20.2 % |
| **Blood pressure (mmHg)** | SBP | 127±19.0 | 121±19.7 | 124±19.4 |
| | DBP | 80±10.8 | 78±11.8 | 79±11.3 |

[a] Continuous variables were expressed as mean ± standard deviations and categorical variables as %
*BMI = Body Mass Index, SBP = Systolic Blood Pressure, DBP = Diastolic Blood Pressure*

| Timepoint 2 | Subgroups | PR (n = 192) | NR (n = 198) | Total (n = 390) |
|---|---|---|---|---|
| **Age (years)** | | 51.5 ± 13.4 | 45.3 ± 13.1 | 48.3±13.6 |
| | Group 1: 19-40 (%) | 34 (17.7) | 76 (38.4) | 28.2 % |
| | Group 2: 41-50 (%) | 45 (23.4) | 39 (19.7) | 21.5 % |
| | Group 3: 51-60 (%) | 50 (26) | 53 (26.8) | 26.4% |
| | Group 4: 61-76 (%) | 63 (32.8) | 30 (15.1) | 23.8 % |
| **Sex (%)** | Female | 103 (53.6) | 108 (54.5) | 54.1 % |
| | Male | 89 (46.4) | 90 (45.5) | 45.9 % |
| **Weight (kg)** | --- | 79±15.5 | 77±15.6 | 78±15.5 |
| **BMI (kg/m²)** | | 28.3 ± 4.6 | 26.6 ± 4.4 | 27.4 ± 4.6 |
| | Group 1: < 18.5 (%) | 1 (0.5) | 5 (2.5) | 1.5 % |
| | Group 2: 18.5-25 (%) | 41 (21.4) | 65 (32.8) | 27.2 % |
| | Group 3: 25-30 (%) | 91 (47.4) | 87 (43.9) | 45.6 % |
| | Group 4: ≥ 30 (%) | 59 (30.7) | 41 (20.7) | 25.6 % |
| **Smoking status (%)** | Non-smoker (%) | 179 (93.2) | 137 (69.2) | 81 % |
| | Smoker (%) | 13 (6.8) | 61 (30.8) | 19 % |
| **Blood pressure (mmHg)** | SBP | 125±17.3 | 120±18.7 | 112.5±18.0 |
| | DBP | 78±10.6 | 78±12.2 | 78±11.4 |

[a] Continuous variables were expressed as mean ± standard deviations and categorical variables as %
*BMI = Body Mass Index; SBP = Systolic Blood Pressure; DBP = Diastolic Blood Pressure*

**Table S3.** FastBio participant medication use.

| | PR | NR | Total (n) |
|---|---|---|---|
| **Blood lipids** | 27 | 13 | 40 |
| **Blood glucose levels** | 7 | 2 | 9 |
| **Osteoporosis** | 6 | 3 | 9 |
| **Thyroid function** | 23 | 26 | 49 |
| **Blood pressure** | 36 | 17 | 53 |
| **Other** | 12 | 21 | 33 |

### REFERENCES

1. Yoo, T.K., et al., *Association between physical activity and insulin resistance using the homeostatic model assessment for insulin resistance independent of waist circumference.* Sci Rep, 2022. **12**(1): p. 6002.
2. Ito, R., et al., *Teneligliptin, a Dipeptidyl Peptidase-4 Inhibitor, Improves Early-Phase Insulin Secretion in Drug-Naive Patients with Type 2 Diabetes.* Drugs R D, 2015. **15**(3): p. 245-51.
3. Geifman, N., R. Cohen, and E. Rubin, *Redefining meaningful age groups in the context of disease.* Age (Dordr), 2013. **35**(6): p. 2357-66.
4. Franssen, T., et al., *Age differences in demographic, social and health-related factors associated with loneliness across the adult life span (19–65 years): a cross-sectional study in the Netherlands.* BMC Public Health, 2020. **20**(1): p. 1118.
5. Swift, H.J., et al., *Categorization by Age*, in *Encyclopedia of Evolutionary Psychological Science*, T.K. Shackelford and V.A. Weekes-Shackelford, Editors. 2018, Springer International Publishing: Cham. p. 1-10.
6. Stenholm, S., et al., *Body mass index as a predictor of healthy and disease-free life expectancy between ages 50 and 75: a multicohort study.* International Journal of Obesity, 2017. **41**(5): p. 769-775.
